# Disease-related baseline cerebrospinal fluid proteomic variation refines biomarker interpretation before CAR-T therapy

**DOI:** 10.64898/2026.08.22.26360883

**Authors:** Tomoko Nomiyama, Daiki Setoyama, Ikumi Yamanaka, Masatoshi Shimo, Kohta Miyawaki, Takuji Yamauchi, Fumiaki Jinnouchi, Teppei Sakoda, Kensuke Sasaki, Hidetaka Nakagaki, Ken Takigawa, Shiho Taniguchi, Takahiro Shima, Yasuo Mori, Sachiko Kanaji, Takahiro A Kato, Yoshikane Kikushige, Koichi Akashi, Yuya Kunisaki, Koji Kato

## Abstract

Pre-infusion cerebrospinal fluid (CSF) proteomics may enable risk stratification for immune effector cell-associated neurotoxicity syndrome (ICANS) after chimeric antigen receptor T-cell therapy, but disease-specific baseline variation may influence biomarker interpretation. We compared pre-infusion CSF proteomic profiles from 28 patients with diffuse large B-cell lymphoma (DLBCL) and 9 with multiple myeloma (MM). Although principal component analysis showed substantial overlap, orthoPLS-DA identified significant disease-associated discrimination supported by permutation testing. Proteins contributing to this separation were enriched for plasma cell-related, extracellular, and metabolic signatures. ICANS occurred in 7 of 28 DLBCL patients but in none of the 9 MM patients. MM cases aligned with the ICANS-negative group in binary analysis while remaining distinct from both DLBCL subgroups in three-group analysis. These findings indicate that pre-infusion CSF proteomics captures disease-specific molecular structure that should be considered when developing and interpreting biomarkers of CAR-T-associated neurotoxicity.

---

Chimeric antigen receptor T-cell (CAR-T) therapy has improved outcomes in hematologic malignancies, but cytokine release syndrome (CRS) and immune effector cell-associated neurotoxicity syndrome (ICANS) remain major toxicities.(Nomiyama *et al*, 2025; Lee *et al*, 2019) A current priority in CAR-T cell therapy is to identify biomarkers of efficacy and toxicity and to develop precision-stratified treatment strategies.(Wang *et al*, 2026) In our previous study,(Nomiyama *et al*, 2025) we examined ICANS risk in patients with diffuse large B-cell lymphoma (DLBCL) and identified high-risk groups before CAR-T therapy using cerebrospinal fluid (CSF) proteomic profiling. However, because ICANS incidence varies according to disease type and target antigen, disease-specific CSF profiles may influence the interpretation of neurotoxicity-associated biomarkers in pre-CAR-T studies. It also remains unclear whether baseline CSF proteomes differ across hematologic malignancies and whether such differences may contribute to disease-specific patterns of ICANS incidence. We therefore compared pretreatment (pre-infusion) CSF proteomic profiles between DLBCL and multiple myeloma (MM) patients undergoing CAR-T therapy.

Pre-infusion CSF samples were available from 37 patients, including 28 with DLBCL and 9 with MM. Post-treatment clinical data, including CRS and ICANS graded according to ASTCT criteria, were collected (**Supplementary Table 1**).(Lee *et al*, 2019) Routine clinical variables, CNS involvement, CSF total protein, and indices of blood contamination were reviewed to assess whether major clinical or pre-analytical factors might account for group-wise proteomic differences. Proteomic data were analyzed by principal component analysis (PCA), PLS-DA, and orthoPLS-DA, with model performance assessed by cross-validation and permutation testing.

Routine clinical characteristics did not clearly distinguish DLBCL from MM. Age, sex, general laboratory variables, CNS involvement, and indices of blood contamination were broadly comparable between groups (**Supplementary Table 1**). CSF total protein remained within the normal range in both groups, arguing against marked barrier dysfunction or substantial blood contamination as the primary explanation for the observed proteomic differences.

We first examined whether unsupervised global proteomic structure separated DLBCL and MM. PCA did not clearly separate the two groups, and the major principal components showed substantial overlap (PC1 17.8%, PC2 11.3%; **Figure 1A**). Thus, disease-associated CSF variation was not among the dominant sources of total variance detectable by unsupervised analysis. Conventional PLS-DA suggested partial discrimination, but model robustness was limited (**Supplementary Figure 1**). In contrast, orthoPLS-DA identified a clearer discriminant structure between DLBCL and MM, with significant permutation support (Q2 = 0.518, empirical p < 0.001; R2Y = 0.951, empirical p = 0.023) (**Figure 1B and 1C**). This result was retained across preprocessing strategies, including autoscaling and Pareto scaling, indicating that the supervised separation was not dependent on a single scaling method (**Supplementary Figure 2**). The lack of separation on PCA, together with the significant orthoPLS-DA result, suggests that disease-associated variation was not a dominant source of total variance but rather a latent discriminative structure masked by broader orthogonal variation.

**Figure 1.**
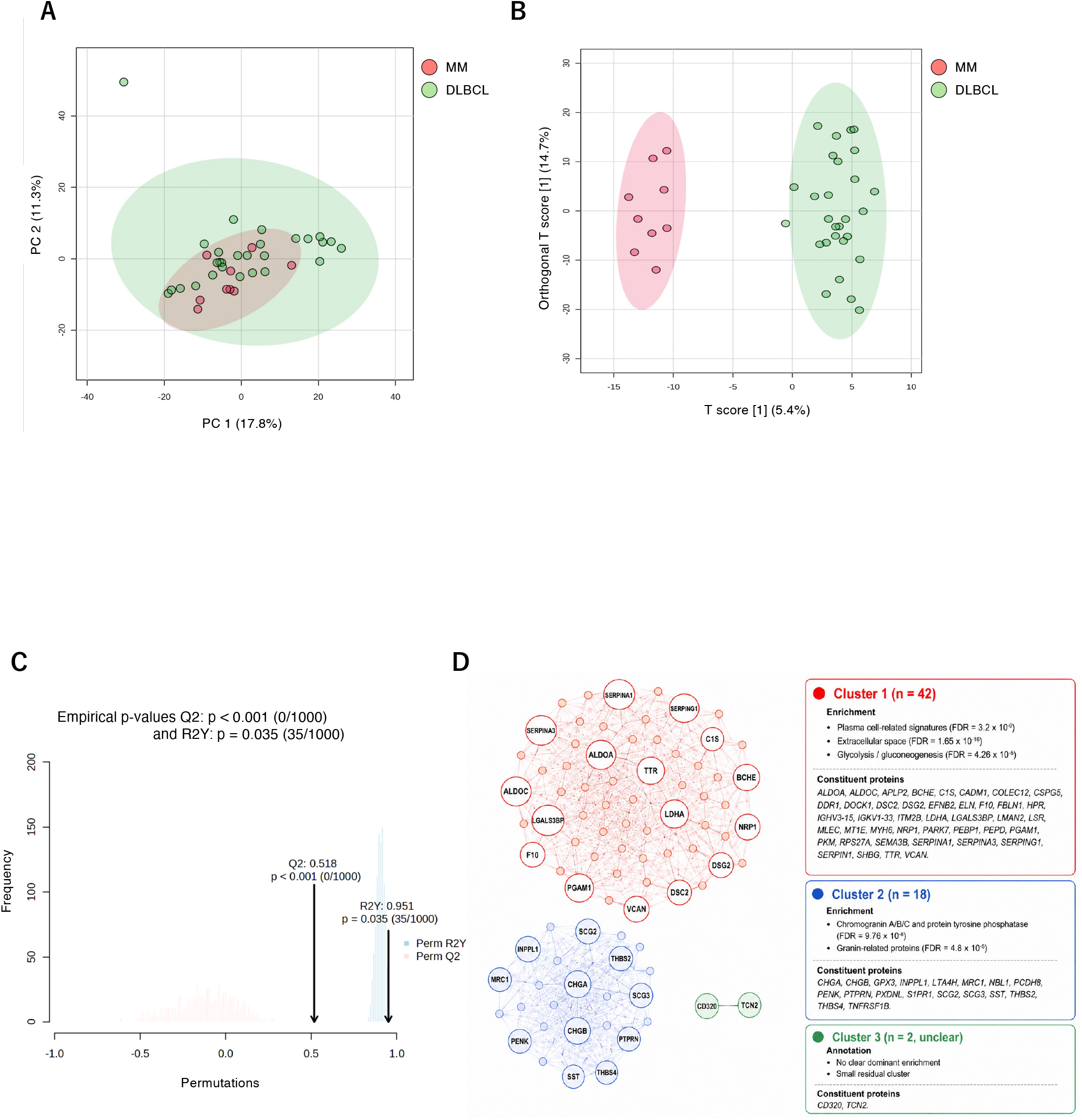
Pre-infusion CSF proteomic structure distinguishes DLBCL and MM. (A) PCA score plot of pre-infusion CSF proteomic profiles from DLBCL and MM. (B) OrthoPLS-DA score plot of pre-infusion CSF proteomic profiles from DLBCL and MM. (C) Permutation test summary for the orthoPLS-DA model. The observed Q2 and R2Y values were significantly greater than those obtained from permuted datasets. (D) STRING network analysis and k-means clustering summary of 120 proteins with VIP > 1.5. Cluster 1 was enriched for plasma cell-related signatures, extracellular space, and glycolysis/gluconeogenesis; Cluster 2 was enriched for chromogranin/granin-related annotations; Cluster 3 showed no clear dominant enrichment.

To explore the biology underlying this discriminant structure, proteins strongly contributing to the orthoPLS-DA model were extracted using VIP >1.5 (n = 120, **Supplementary Table 2**) and subjected to STRING network analysis followed by k-means clustering (k = 3) (**Figure 1D**). The largest cluster (42 proteins) showed highly significant enrichment for plasma cell-related signatures (FDR = 3.2 × 10^−9^), extracellular space (FDR = 1.65 × 10^−16^), and glycolysis/gluconeogenesis (FDR = 4.26 × 10^−5^). A second cluster (21 proteins) was enriched for annotations related to Chromogranin A/B/C and protein tyrosine phosphatase (FDR = 9.76 × 10^−8^), as well as granin-related proteins (FDR = 4.8 × 10^−5^) (**Supplementary Table 3**). These findings support the biological plausibility of the supervised discriminant structure and suggest that disease-associated baseline CSF variation reflects a composite extracellular proteomic signature with a strong plasma cell-associated component. Together, these results indicate that pre-infusion CSF proteomics can capture disease-associated molecular structure beyond clinical phenotyping.(Johnson *et al*, 2023; Rade *et al*, 2026)

We next examined the relationship between baseline proteomic structure and subsequent ICANS. ICANS was not observed in any MM patient in this cohort (0/9), whereas it occurred in 7 of 28 DLBCL patients. When patients were dichotomized according to subsequent ICANS status, all MM patients were classified within the ICANS-negative rather than the ICANS-positive group (**Figure 2A**). Thus, the MM cohort was not only clinically free of ICANS, but was also assigned entirely to the non-ICANS side of the discriminant structure. However, when three-group discrimination was performed using DLBCL with ICANS, DLBCL without ICANS, and MM, the MM cases formed a distinct cluster, while the two DLBCL subgroups occupied an intermediate position (**Figure 2B**). These findings suggest that disease-associated baseline structure and ICANS-associated variation are partially overlapping but non-identical axes (**Figure 2C**). Specifically, MM aligned with the non-ICANS direction in binary analysis, yet retained a distinct disease-specific baseline signature in three-group analysis. This observation raises the possibility that some baseline molecular features may be associated with reduced susceptibility to ICANS across disease contexts, although this interpretation remains exploratory and will require validation in larger multi-disease cohorts. This interpretation is consistent with the lower prominence of ICANS in BCMA-directed CAR-T therapy for MM compared with CD19-directed CAR-T therapy for lymphoma, while acknowledging that clinically relevant neurologic events may still occur in selected MM settings.(Cohen *et al*, 2022; Gaballa *et al*, 2025) Recent studies have further supported the value of molecular profiling for toxicity stratification in CD19- and BCMA-directed CAR-T therapy.(Rade *et al*, 2026; Gomez-Llobell *et al*, 2025; Zugasti *et al*, 2025)

**Figure 2.**
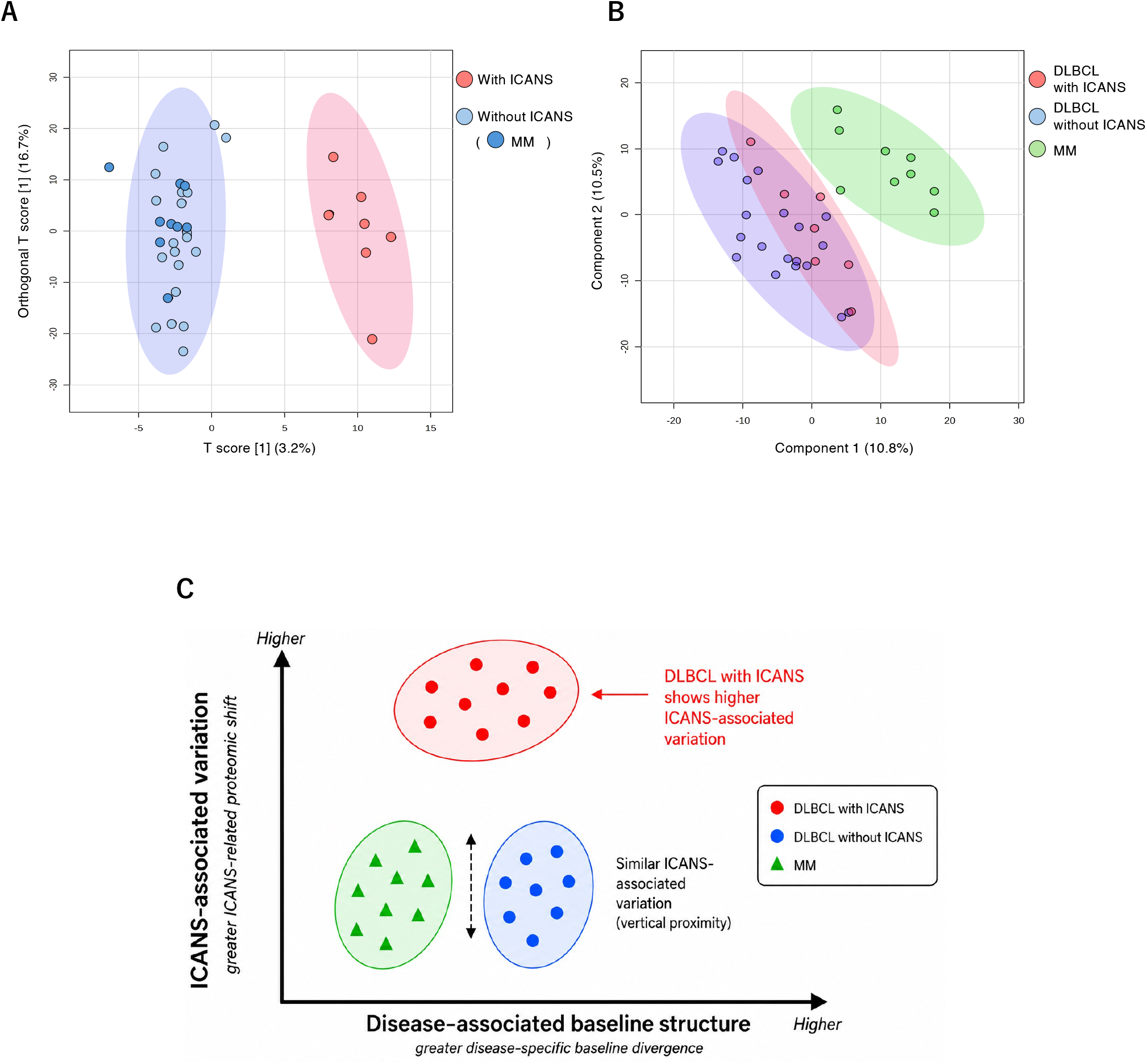
Relationship between baseline CSF proteomic structure and subsequent ICANS. (A) Binary discriminant analysis according to subsequent ICANS status. All MM cases were classified within the ICANS-negative group. (B) Three-group discriminant analysis including DLBCL with ICANS, DLBCL without ICANS, and MM. MM formed a distinct cluster, while the two DLBCL subgroups occupied an intermediate position. (C) Conceptual summary schema illustrating the relationship between disease-associated baseline structure and ICANS-associated variation. MM aligns with the non-ICANS direction in binary analysis yet remains distinct from both DLBCL subgroups in three-group analysis, supporting the interpretation that pre-infusion CSF proteomics captures both malignancy-related baseline context and variation associated with subsequent neurotoxicity risk.

Together, these findings provide important implications for the development of pre-CAR-T CSF biomarker models. Our data suggest that pre-infusion CSF proteomics can capture variation associated with subsequent ICANS risk, as well as biologically significant baseline patterns linked to the underlying malignancy. Disease stratification is therefore likely to improve both biological interpretation and predictive modeling in future studies of CAR-T-associated neurotoxicity. This point refines, rather than weakens, the implications of our previous report(Nomiyama *et al*, 2025): pre-infusion CSF proteomics remains a promising biomarker platform, but its interpretation should account for disease-specific molecular context.(Wang *et al*, 2026)

This study has limitations. It was a single-center exploratory analysis with a limited number of MM cases. Disease separation was not apparent on PCA and required supervised modeling, indicating that the relevant signal is subtle rather than dominant. Although orthoPLS-DA showed permutation-supported discrimination across scaling approaches, external validation remains necessary. In addition, the ICANS-related analyses were exploratory and should not be interpreted as a definitive predictive model.

In summary, pre-infusion CSF proteomics revealed disease-related baseline structure across DLBCL and MM that may influence interpretation of pre-CAR-T biomarker signatures. MM patients were uniformly free of ICANS, aligned with the ICANS-negative group in binary analysis, yet remained distinct in three-group analysis. These findings support pre-infusion CSF proteomics as a biomarker platform while underscoring the need to account for disease-specific baseline variation.

## Acknowledgments

This study was supported by the Japan Agency for Medical Research and Development (AMED; grant number 26ama221450h0002 to K.K., D.S., and Y.K.), the Japan Society for the Promotion of Science (JSPS KAKENHI; grant number 25K19578 to T.N.), and a Charitable Trust Laboratory Medicine Research Foundation of Japan to T.N. We would like to thank Arisa Matsuyama and Ayumi Inayoshi for their assistance with the research, Kyohei Mori and Miyuki Yoshikawa for their management and provision of clinical information.

## Data Availability

The proteomics data generated in this study are available from the corresponding author upon reasonable request.

## Contributions

TN, DS and KK designed the study; TN and IY collected the clinical samples and patients’ data; TN and DS collected and analyzed the data; TN, DS, and KK interpreted the data; TN and DS drafted the manuscript; DS and KK finalized the manuscript. IY, MS, KM, TY, FJ, TS, KS, HN, KT, ST, TS, YM, SK, TK, Y.Kikushige, KA, and Y.Kunisaki provided clinical feedback. All authors approved the manuscript.

## Ethics approval and consent to participate

This study was a retrospective analysis conducted in accordance with the principles outlined in the Declaration of Helsinki and was approved by the Ethics Committee of Kyushu University (IRB numbers: 22213 and 23399). Informed consent was obtained using an opt-out methodology, whereby study details were disclosed on the institutional website, allowing participants the opportunity to decline participation. Data and CSF samples from individuals who did not opt out were included in the analysis. All collected data were anonymized prior to analysis to safeguard the privacy of participants.

## Supplementary information

### Materials and methods

#### CSF sample collection and proteomic analysis

CSF sample collection, protein digestion, LC-MS/MS analysis, and DIA-based protein identification were performed essentially as previously described in our recent *Leukemia* report. Briefly, residual CSF samples obtained before CAR-T cell infusion were centrifuged, and the supernatants were stored at ™80°C until analysis. CSF proteins were digested using the Rapid-Digestion Trypsin/Lys-C Kit (Promega, #VA1061), and proteomic analysis was conducted on the EVOSEP ONE/Q-Exactive platform using data-independent acquisition (DIA). DIA data were analyzed using DIA-NN software (version 1.9.1) with reference to a human protein FASTA database.

#### Multivariate and network analyses

Multivariate data analysis, including principal component analysis (PCA), clustering heat map analysis, PLS-DA, and orthoPLS-DA, was performed using MetaboAnalyst 6.0. STRING version 12.0 was used for protein network analysis, and k-means clustering was applied to proteins strongly contributing to the orthoPLS-DA model. Patient demographics were summarized using the R package *gtsummary*.

**Supplementary Table 1.** Clinical characteristics of patients with DLBCL and MM treated with CAR-T therapy. Baseline clinical and laboratory characteristics of the study cohort, including age, sex, routine clinical variables, ICANS status, CSF total protein, and indices of blood contamination, are compared between diffuse large B-cell lymphoma (DLBCL) and multiple myeloma (MM).

| Characteristic | Overall N = 36<sup>1</sup> | DLBCL N = 27<sup>1</sup> | MM N = 9<sup>1</sup> | p-value<sup>2</sup> |
| --- | --- | --- | --- | --- |
| Age | 62 (48, 72) | 62 (53, 72) | 61 (48, 65) | 0.3 |
| Sex (M/F) | 19/17 | 14/13 | 5/4 | >0.9 |
| ICANS; |  |  |  |  |
| Negative | 29 (81%) | 20 (74%) | 9 (100%) |  |
| Positive | 7 (19%) | 7 (26%) | 0 (0%) |  |
| Total Protein | 45 (29, 52) | 48 (30, 53) | 40 (29, 44) | 0.11 |
| Glu | 62 (55, 72) | 62 (54, 69) | 67 (59, 76) | 0.2 |
| Na | 149.5 (147.0, 151.5) | 149.0 (147.0, 151.0) | 150.0 (148.0, 152.0) | 0.6 |
| K | 2.90 (2.90, 3.00) | 3.00 (2.90, 3.10) | 2.90 (2.80, 3.00) | 0.14 |
| LDH | 21.0 (17.0, 25.0) | 20.5 (19.0, 26.0) | 21.0 (15.0, 24.0) | 0.3 |
| WBC | 2.00 (1.00, 2.00) | 2.00 (1.00, 3.00) | 1.00 (1.00, 2.00) | 0.7 |

DLBCL, Diffuse large B-cell lymphoma
Glu, glucose; LDH, lactate dehydrogenase; WBC, white blood cell
<sup>1</sup> Median (minimun, maximun); Mean(SD); n (%)<sup>2</sup> Welch Two Sample t-test; Fisher's exact test

**Supplementary Table 2.**
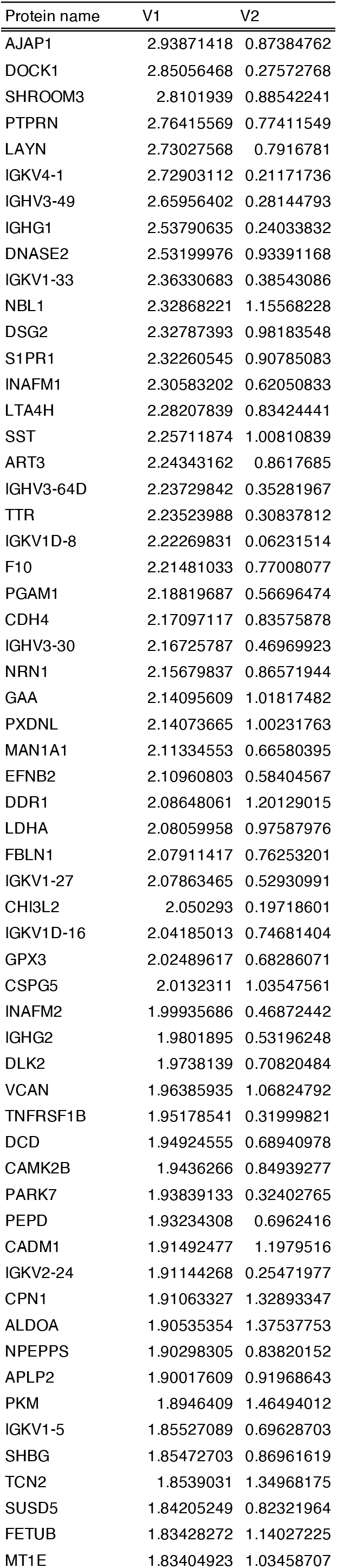
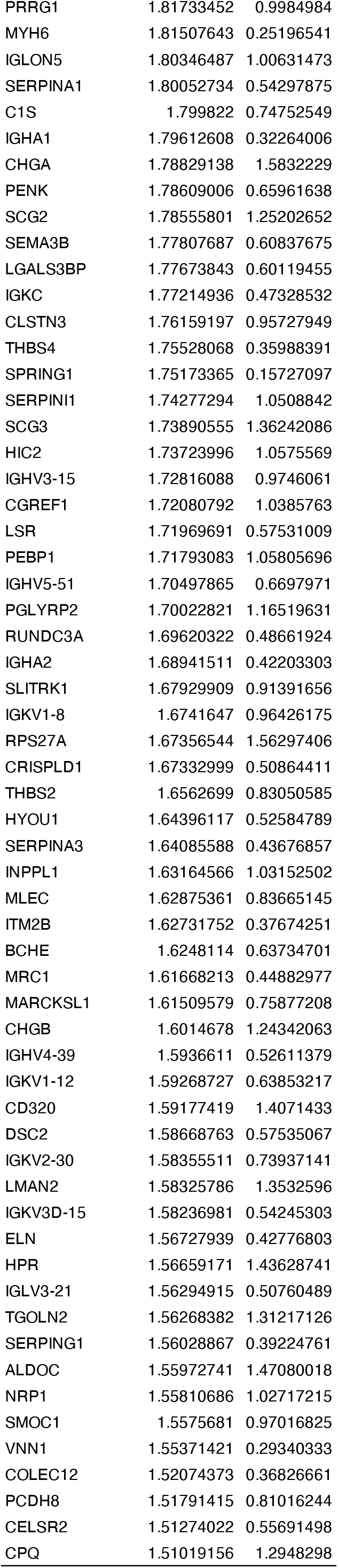
Proteins with VIP > 1.5 in the orthoPLS-DA model. List of proteins contributing strongly to the orthoPLS-DA-based discrimination between DLBCL and MM, defined by a variable importance in projection (VIP) score > 1.5. Protein names, gene symbols, and, where applicable, cluster assignments are shown.

| Protein name | V1 | V2 |
| --- | --- | --- |
| AJAP1 | 2.93871418 | 0.87384762 |
| DOCK1 | 2.85056468 | 0.27572768 |
| SHROOM3 | 2.8101939 | 0.88542241 |
| PTPRN | 2.76415569 | 0.77411549 |
| LAYN | 2.73027568 | 0.7916781 |
| IGKV4-1 | 2.72903112 | 0.21171736 |
| IGHV3-49 | 2.65956402 | 0.28144793 |
| IGHG1 | 2.53790635 | 0.24033832 |
| DNASE2 | 2.53199976 | 0.93391168 |
| IGKV1-33 | 2.36330683 | 0.38543086 |
| NBL1 | 2.32868221 | 1.15568228 |
| DSG2 | 2.32787393 | 0.98183548 |
| S1PR1 | 2.32260545 | 0.90785083 |
| INAFM1 | 2.30583202 | 0.62050833 |
| LTA4H | 2.28207839 | 0.83424441 |
| SST | 2.25711874 | 1.00810839 |
| ART3 | 2.24343162 | 0.8617685 |
| IGHV3-64D | 2.23729842 | 0.35281967 |
| TTR | 2.23523988 | 0.30837812 |
| IGKV1D-8 | 2.22269831 | 0.06231514 |
| F10 | 2.21481033 | 0.77008077 |
| PGAM1 | 2.18819687 | 0.56696474 |
| CDH4 | 2.17097117 | 0.83575878 |
| IGHV3-30 | 2.16725787 | 0.46969923 |
| NRN1 | 2.15679837 | 0.86571944 |
| GAA | 2.14095609 | 1.01817482 |
| PXDNL | 2.14073665 | 1.00231763 |
| MAN1A1 | 2.11334553 | 0.66580395 |
| EFNB2 | 2.10960803 | 0.58404567 |
| DDR1 | 2.08648061 | 1.20129015 |
| LDHA | 2.08059958 | 0.97587976 |
| FBLN1 | 2.07911417 | 0.76253201 |
| IGKV1-27 | 2.07863465 | 0.52930991 |
| CHI3L2 | 2.050293 | 0.19718601 |
| IGKV1D-16 | 2.04185013 | 0.74681404 |
| GPX3 | 2.02489617 | 0.68286071 |
| CSPG5 | 2.0132311 | 1.03547561 |
| INAFM2 | 1.99935686 | 0.46872442 |
| IGHG2 | 1.9801895 | 0.53196248 |
| DLK2 | 1.9738139 | 0.70820484 |
| VCAN | 1.96385935 | 1.06824792 |
| TNFRSF1B | 1.95178541 | 0.31999821 |
| DCD | 1.94924555 | 0.68940978 |
| CAMK2B | 1.9436266 | 0.84939277 |
| PARK7 | 1.93839133 | 0.32402765 |
| PEPD | 1.93234308 | 0.6962416 |
| CADM1 | 1.91492477 | 1.1979516 |
| IGKV2-24 | 1.91144268 | 0.25471977 |
| CPN1 | 1.91063327 | 1.32893347 |
| ALDOA | 1.90535354 | 1.37537753 |
| NPEPPS | 1.90298305 | 0.83820152 |
| APLP2 | 1.90017609 | 0.91968643 |
| PKM | 1.8946409 | 1.46494012 |
| IGKV1-5 | 1.85527089 | 0.69628703 |
| SHBG | 1.85472703 | 0.86961619 |
| TCN2 | 1.8539031 | 1.34968175 |
| SUSD5 | 1.84205249 | 0.82321964 |
| FETUB | 1.83428272 | 1.14027225 |
| MT1E | 1.83404923 | 1.03458707 |
| PRRG1 | 1.81733452 | 0.9984984 |
| MYH6 | 1.81507643 | 0.25196541 |
| IGLON5 | 1.80346487 | 1.00631473 |
| SERPINA1 | 1.80052734 | 0.54297875 |
| C1S | 1.799822 | 0.74752549 |
| IGHA1 | 1.79612608 | 0.32264006 |
| CHGA | 1.78829138 | 1.5832229 |
| PENK | 1.78609006 | 0.65961638 |
| SCG2 | 1.78555801 | 1.25202652 |
| SEMA3B | 1.77807687 | 0.60837675 |
| LGALS3BP | 1.77673843 | 0.60119455 |
| IGKC | 1.77214936 | 0.47328532 |
| CLSTN3 | 1.76159197 | 0.95727949 |
| THBS4 | 1.75528068 | 0.35988391 |
| SPRING1 | 1.75173365 | 0.15727097 |
| SERPINI1 | 1.74277294 | 1.0508842 |
| SCG3 | 1.73890555 | 1.36242086 |
| HIC2 | 1.73723996 | 1.0575569 |
| IGHV3-15 | 1.72816088 | 0.9746061 |
| CGREF1 | 1.72080792 | 1.0385763 |
| LSR | 1.71969691 | 0.57531009 |
| PEBP1 | 1.71793083 | 1.05805696 |
| IGHV5-51 | 1.70497865 | 0.6697971 |
| PGLYRP2 | 1.70022821 | 1.16519631 |
| RUNDC3A | 1.69620322 | 0.48661924 |
| IGHA2 | 1.68941511 | 0.42203303 |
| SLITRK1 | 1.67929909 | 0.91391656 |
| IGKV1-8 | 1.6741647 | 0.96426175 |
| RPS27A | 1.67356544 | 1.56297406 |
| CRISPLD1 | 1.67332999 | 0.50864411 |
| THBS2 | 1.6562699 | 0.83050585 |
| HYOU1 | 1.64396117 | 0.52584789 |
| SERPINA3 | 1.64085588 | 0.43676857 |
| INPPL1 | 1.63164566 | 1.03152502 |
| MLEC | 1.62875361 | 0.83665145 |
| ITM2B | 1.62731752 | 0.37674251 |
| BCHE | 1.6248114 | 0.63734701 |
| MRC1 | 1.61668213 | 0.44882977 |
| MARCKSL1 | 1.61509579 | 0.75877208 |
| CHGB | 1.6014678 | 1.24342063 |
| IGHV4-39 | 1.5936611 | 0.52611379 |
| IGKV1-12 | 1.59268727 | 0.63853217 |
| CD320 | 1.59177419 | 1.4071433 |
| DSC2 | 1.58668763 | 0.57535067 |
| IGKV2-30 | 1.58355511 | 0.73937141 |
| LMAN2 | 1.58325786 | 1.3532596 |
| IGKV3D-15 | 1.58236981 | 0.54245303 |
| ELN | 1.56727939 | 0.42776803 |
| HPR | 1.56659171 | 1.43628741 |
| IGLV3-21 | 1.56294915 | 0.50760489 |
| TGOLN2 | 1.56268382 | 1.31217126 |
| SERPING1 | 1.56028867 | 0.39224761 |
| ALDOC | 1.55972741 | 1.47080018 |
| NRP1 | 1.55810686 | 1.02717215 |
| SMOC1 | 1.5575681 | 0.97016825 |
| VNN1 | 1.55371421 | 0.29340333 |
| COLEC12 | 1.52074373 | 0.36826661 |
| PCDH8 | 1.51791415 | 0.81016244 |
| CELSR2 | 1.51274022 | 0.55691498 |
| CPQ | 1.51019156 | 1.2948298 |

**Supplementary Table 3.**
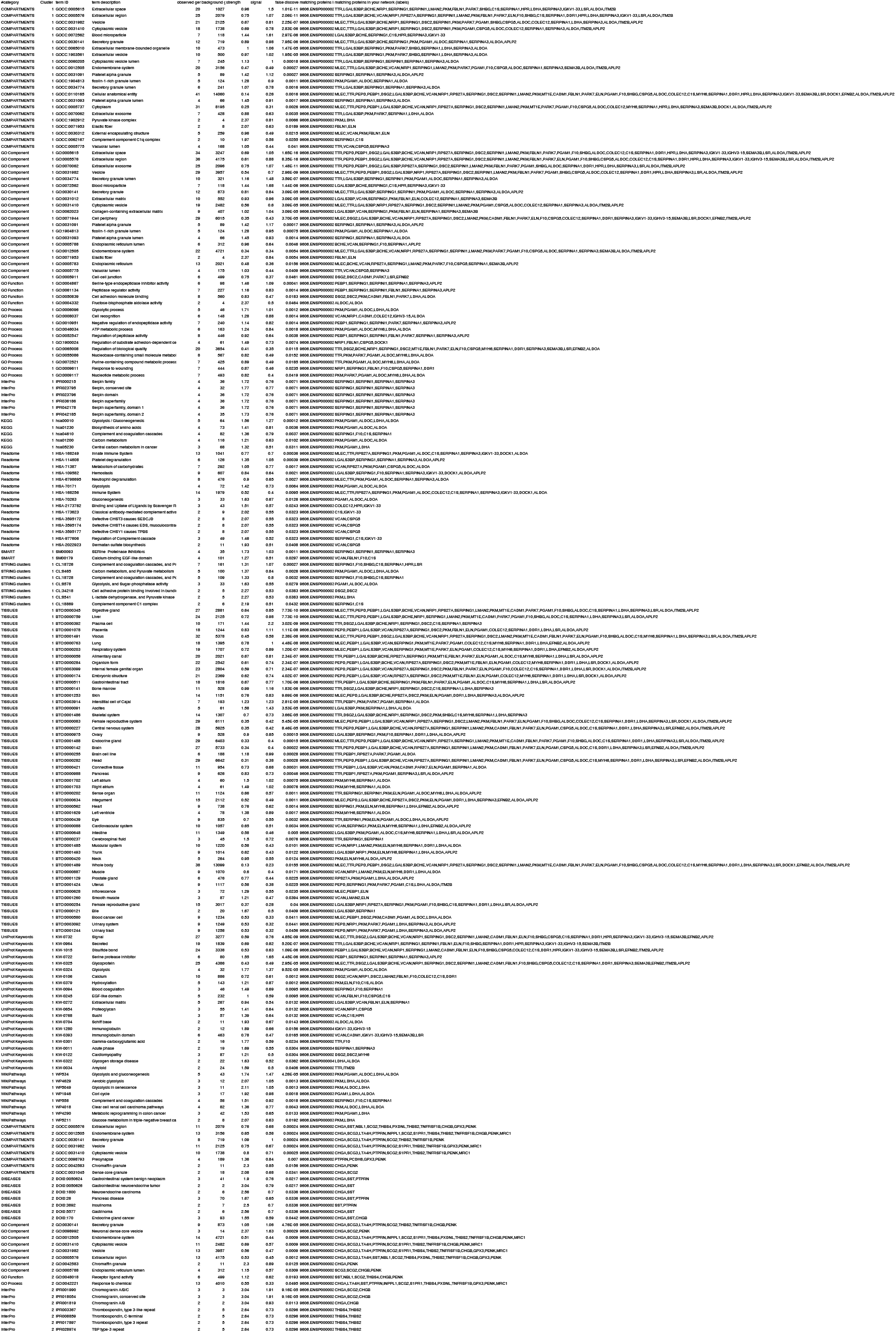
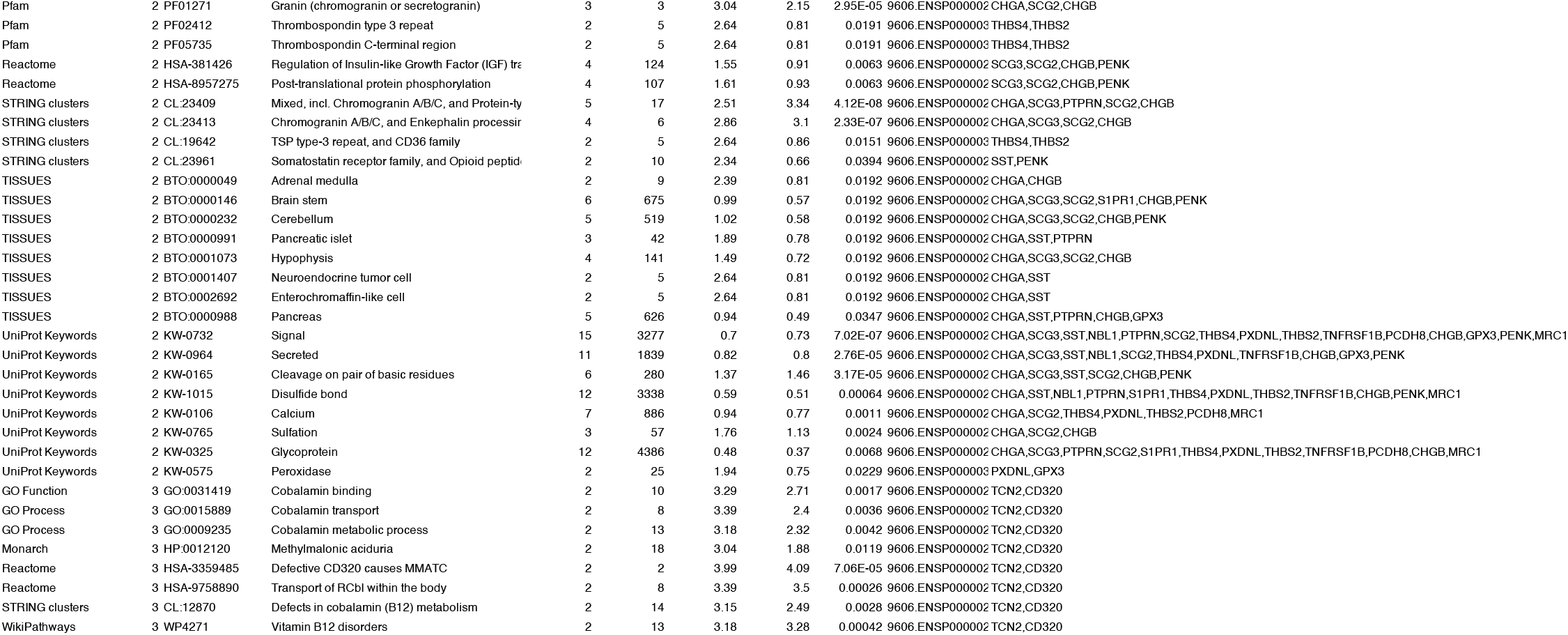
Cluster enrichment analysis of proteins contributing to the orthoPLS-DA model. Detailed enrichment results for k-means clusters derived from STRING network analysis of proteins with VIP > 1.5. Enriched annotations, corresponding false discovery rate (FDR) values, and constituent proteins are shown for each cluster.

**Supplementary Figure 1.**
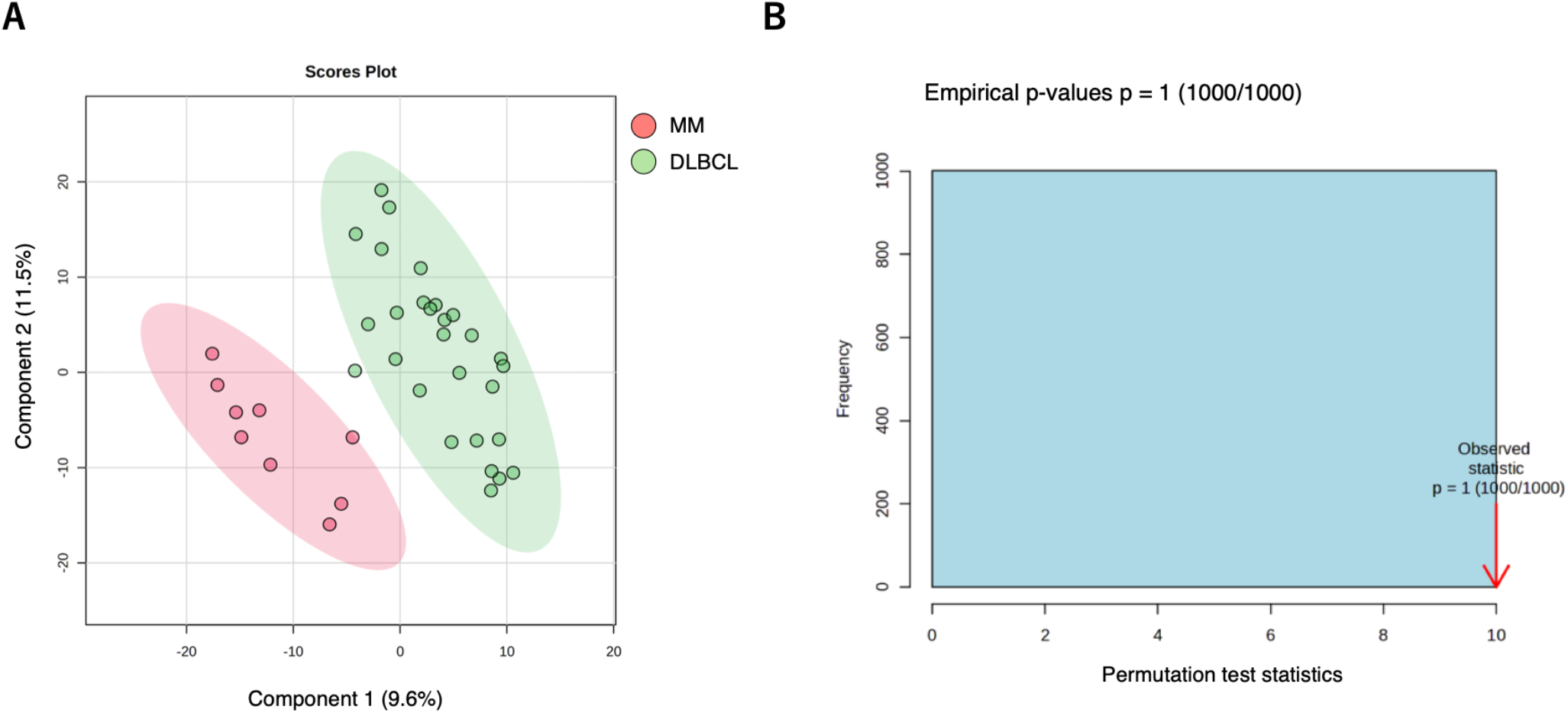
Conventional PLS-DA showed limited robustness. (A) Score plot. (B) Permutation test summary showing limited model robustness.

**Supplementary Figure 2.**
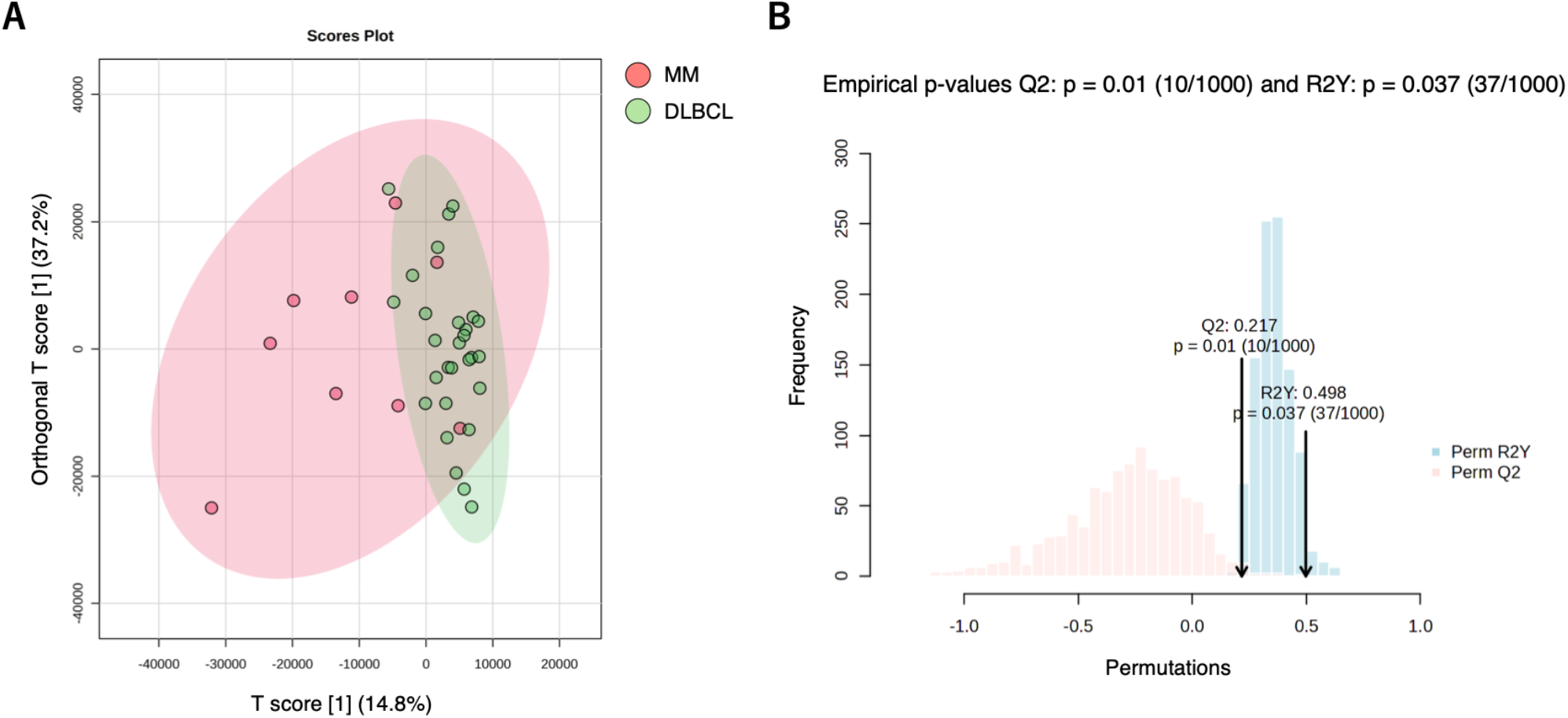
Sensitivity analysis under Pareto scaling. (A) OrthoPLS-DA score plot under Pareto scaling. (B) Permutation test summary showing retained significance under an alternative preprocessing strategy.

## References

Cohen, A.D., Parekh, S., Santomasso, B.D., Gállego Pérez-Larraya, J., Van De Donk, N.W.C.J., Arnulf, B., Mateos, M.-V., Lendvai, N., Jackson, C.C., De Braganca, K.C., Schecter, J.M., Marquez, L., Lee, E., Cornax, I., Zudaire, E., Li, C., Olyslager, Y., Madduri, D., Varsos, H., Pacaud, L., et al (2022) Incidence and management of CAR-T neurotoxicity in patients with multiple myeloma treated with ciltacabtagene autoleucel in CARTITUDE studies. Blood Cancer Journal, 12, 32.

Gaballa, M.R., Puglianini, O.C., Cohen, A., Vogl, D., Chung, A., Ferreri, C.J., Voorhees, P., Hansen, D.K. & Patel, K.K. (2025) BCMA-directed CAR T-cell therapy in patients with multiple myeloma and CNS involvement. Blood Advances, 9, 1171–1180.

Gomez-Llobell, M., Serra Smith, C., Gómez-Costas, D., Gómez-Centurión, I., García Domínguez, J.M., Escudero-Vilaplana, V., Fernández Bullido, Y., Carbonell, D., Pion, M., Bailén, R., Gil-Perotin, S., Martínez Ginés, M.L., Revuelta-Herrero, J.L., Fernández-Caldas, P., Fernández, V.A.P., Collado-Borrell, R., Bastos-Oreiro, M., Villanueva-Bueno, C., García-Sanz, R. & Kwon, M. (2025) ICANS risk model in CD19 CAR-T therapy: insights from serum and CSF cytokine profiling. Bone Marrow Transplantation, 60, 1351–1360.

Johnson, E.C.B., Bian, S., Haque, R.U., Carter, E.K., Watson, C.M., Gordon, B.A., Ping, L., Duong, D.M., Epstein, M.P., McDade, E., Barthélemy, N.R., Karch, C.M., Xiong, C., Cruchaga, C., Perrin, R.J., Wingo, A.P., Wingo, T.S., Chhatwal, J.P., Day, G.S., Noble, J.M., et al (2023) Cerebrospinal fluid proteomics define the natural history of autosomal dominant Alzheimer’s disease. Nature Medicine, 29, 1979–1988.

Lee, D.W., Santomasso, B.D., Locke, F.L., Ghobadi, A., Turtle, C.J., Brudno, J.N., Maus, M.V., Park, J.H., Mead, E., Pavletic, S., Go, W.Y., Eldjerou, L., Gardner, R.A., Frey, N., Curran, K.J., Peggs, K., Pasquini, M., DiPersio, J.F., Van Den Brink, M.R.M., Komanduri, K.V., et al (2019) ASTCT Consensus Grading for Cytokine Release Syndrome and Neurologic Toxicity Associated with Immune Effector Cells. Biology of Blood and Marrow Transplantation, 25, 625–638.

Nomiyama, T., Setoyama, D., Yamanaka, I., Shimo, M., Miyawaki, K., Yamauchi, T., Jinnouchi, F., Sakoda, T., Sasaki, K., Shima, T., Kikushige, Y., Mori, Y., Akashi, K., Kato, K. & Kunisaki, Y. (2025) Cerebrospinal fluid proteomics exerts predictive potential for immune effector cell-associated neurotoxicity syndrome (ICANS) in CAR-T cell therapy. Leukemia, 39, 983–987.

Rade, M., Fandrei, D., Kreuz, M., Seiffert, S., Grahnert, A., Friedrich, M., Wiemers, T., Born, P., Fischer, L., Weidner, H., Hofbauer, L.C., Baber, R., Wang, S.Y., Bach, E., Hoffmann, S., Scolnick, J., Friedrich, M., Keramati, F., Brazda, P., Sebestyen, Z., et al (2026) A longitudinal single-cell atlas to predict outcome and toxicity after BCMA-directed CAR T cell therapy in multiple myeloma. Cancer Cell, 44, 586–603.e9.

Wang, Y., Cheng, W., Kang, K., Niu, T., Zhao, A. & Wu, Y. (2026) Biomarker-empowered precision navigation of CAR-T cell therapy. Molecular Cancer Available at: https://link.springer.com/10.1186/s12943-026-02647-0 [Accessed May 21, 2026].

Zugasti, I., Espinosa-Aroca, Lady, Fidyt, K., Mulens-Arias, V., Diaz-Beya, M., Juan, M., Urbano-Ispizua, Á., Esteve, J., Velasco-Hernandez, T. & Menéndez, P. (2025) CAR-T cell therapy for cancer: current challenges and future directions. Signal Transduction and Targeted Therapy, 10, 210.

